# Modeling remdesivir antiviral efficacy in COVID-19 hospitalized patients of the randomized, controlled, open-label DisCoVeRy trial

**DOI:** 10.1101/2021.10.19.21265209

**Authors:** Guillaume Lingas, Nadège Néant, Alexandre Gaymard, Drifa Belhadi, Gilles Peytavin, Maya Hites, Thérèse Staub, Richard Greil, Jose-Artur Paiva, Julien Poissy, Nathan Peiffer-Smadja, Dominique Costagliola, Yazdan Yazdanpanah, Florent Wallet, Amandine Gagneux-Brunon, France Mentré, Florence Ader, Charles Burdet, Jérémie Guedj, Maude Bouscambert-Duchamp, the DisCoVeRy study group

## Abstract

Despite several clinical studies, the antiviral efficacy of remdesivir in COVID-19 hospitalized patients remains controversial. We analyzed nasopharyngeal normalized viral loads collected in the 29 days following randomization from 665 hospitalized patients included in the DisCoVeRy trial, allocated to either standard of care (SoC, N=329) or SoC + remdesivir for 10 days (N=336). We used a mathematical model to reconstruct viral kinetic profiles and estimate the antiviral efficacy of remdesivir in reducing viral production. To identify factors associated with viral kinetics, additional analyses were conducted stratified either on time of treatment initiation (≤ or > 7 days since symptom onset) or viral load at randomization (< or ≥ 3.5 log_10_ copies/10^4^ cells). In our model, remdesivir reduced viral production by 2-fold on average (95%CI: 1.5-3.2). Using the estimated parameter of the model, simulations predict that remdesivir reduces time to viral clearance by 0.7 day compared to SoC, with large inter-individual variabilities (Inter-Quartile Range, IQR: 0.0-1.3 days). Exploratory analyses suggest that remdesivir had a larger impact in patients with a high viral load at randomization, reducing viral production by 5-fold on average (95%CI: 2.8-25), leading to a predicted median reduction in the time to viral clearance of 2.4 days (IQR: 0.9-4.5 days).

In summary, our model shows that remdesivir reduces viral production from infected cells by a factor 2, leading to a median reduction of 0.7 days in the time to viral clearance compared to SoC. The efficacy was larger in patients with high level of viral load at treatment initiation.

**One sentence summary:** Remdesivir reduces the time to SARS-CoV-2 clearance by 1 day in hospitalized patients, and up to 3 days in those with high viral load at admission.

## Introduction

Remarkable progresses have recently been made in finding effective antiviral treatments in SARS-CoV-2 infected patients that can prevent disease progression and hospitalization *(1–3)*. However, these treatments need to be administered early in the infection, typically in the first week after symptom onset, to be fully effective *(1–3)*. Although encouraging results have been found in the Recovery trial *(4)*, the role of antiviral treatments in hospitalized patients to prevent mechanical ventilation and death remains unclear. Among these drugs, remdesivir has received an emergency use authorization for the treatment of COVID-19 hospitalized patients in several countries, and is approved for hospitalized patients in the United States *(5)*. However its clinical efficacy remains controversial, with some randomized clinical trials pointing to an efficacy in preventing disease worsening, and others finding no efficacy *(6–9)*.

Remdesivir is a nucleoside analogue prodrug inhibiting RNA polymerase activity of several viruses *(10)*, that has shown antiviral activity against SARS-CoV-2 both *in vitro* and in animal models *(11–13)*. An important element to precisely evaluate remdesivir is to analyze its effect on viral dynamics, which is likely a prerequisite to clinical efficacy. However, results from the literature are still scarce. In three randomized and controlled clinical trials, no difference in viral load levels were found between hospitalized patients receiving remdesivir and those that did not receive remdesivir *(6, 9, 14)*. In two of these studies, the analysis of the effect was hampered by the limited number of patients (with 237 and 181 patients included in *(6, 9)*) and by the design of the analysis that compared viral load at different time points and not the overall effect on viral dynamics *(6)*. Moreover, results interpretation remain limited by the heterogeneity in the biological samples and the molecular techniques used. In the larger DisCoVeRy trial that used normalized viral loads *(14)*, no effects of remdesivir on viral dynamics was found, but the variability of the time interval between symptoms onset and treatment initiation, which is a key factor for antiviral drug evaluation *(15, 16)*, was not considered. One potential approach to address this issue is to use a model-based approach to reconstitute the precise effect of treatment on the course of viral dynamics, as previously shown in a cohort of hospitalized patients *(17)*.

Here, we developed this approach to estimate the effect of remdesivir in inhibiting viral replication. We used normalized and centralized data on viral kinetics from the European randomized controlled DisCoVeRy trial that compared the efficacy of remdesivir plus standard of care (SoC) to SoC alone in COVID-19 hospitalized patients*(14, 18)*.

## Results

### Baseline characteristics and viral load data of analyzed population

A total of 832 patients were evaluable for the primary intention-to-treat analysis *(19)*, among whom 684 had at least one nasopharyngeal viral load available (Fig. 1. & Supplementary Fig. S1.). Patients randomized in the remdesivir arm who did not receive treatment were excluded from the present analysis (N=2). A total of 17 patients who were randomized more than 20 days after symptoms onset were also removed from analysis (see Methods), leaving a total of 665 patients (SoC alone, N=329; SoC + remdesivir, N=336) included in the present analysis (Supplementary Figs. S1 & S2). The median delay between symptom onset and randomization was 9 days (Interquartile range, IQR = 7-11 days, Table 1.). Patients were mostly male (N=457, 68.7%), younger than 65-year-old (N=349, 52.5%) and randomized more than 7 days after symptom onset (N=455, 68.4%). A median of 4 nasopharyngeal swabs were available per patient (IQR=3-6), and the median viral load at randomization was 3.2 log_10_ copies/10^4^ cells.

**Fig. 1.**
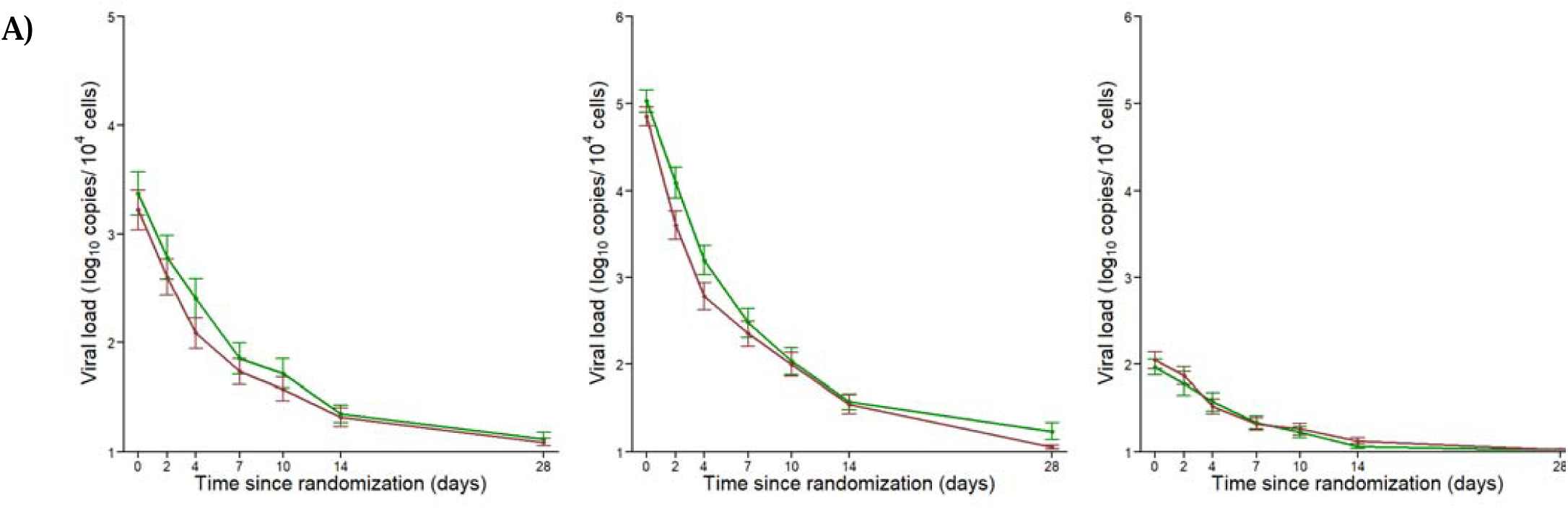
Nasopharyngeal normalized viral load data in 665 patients from DisCoVeRy trial analyzed in the present study. A) SARS-CoV-2 nasopharyngeal viral load according to the time since randomization. B) and C) SARS-CoV-2 nasopharyngeal viral load according to the time since randomization in patients 3.5 log_10_ copies/10^4^ cells (N=184, B) and <3.5 log_10_ copies/10^4^ cells (N=236, C). Data are presented as means (95%CI). Red: patients receiving remdesivir + SoC. Green: patients receiving SoC only.

**Table 1.** Baseline characteristics of the analyzed population at randomization.

| Characteristics | Standard of care |  | Standard of care + Remdesivir |  |
| --- | --- | --- | --- | --- |
|  | (N=329) |  | (N=336) |  |
|  | Median (IQR) or<br>n (%) | Missing data (%) | Median (IQR)<br>or n (%) | Missing data (%) |
| **Male gender** | 222 (67.5%) | 0 (0%) | 235 (69.9%) | 0 (0%) |
| **Age (years)** | 64 (53-72) | 0 (0%) | 63 (55-73) | 0 (0%) |
| **<65** | 169 (51.4%) | 0 (0%) | 180 (53.6%) | 0 (0%) |
| **≥65** | 160 (48.6%) | 0 (0%) | 156 (46.4%) | 0 (0%) |
| **Time since symptom onset (days)** | 9 (7-11) | 5 (1.5%) | 9 (7-11) | 6 (1.8%) |
| **Normalized viral load at**<br>**randomization (log<sub>10</sub> copies/10<sup>4</sup> cells)** | 3.2 (1.9-4.5) | 131 (39.8%) | 3.2 (1.8-4.5) | 114 (33.9%) |

### Viral dynamics parameters

We used a target-cell limited model with an effect of age on the loss rate of infected cells, δ, (see Methods) which was estimated to 0.88 d^-1^ (95%CI: 0.80-0.96) in individuals aged < 65-year-old and 0.75 d^-1^ (95%CI: 0.67-0.84) in those ≥ 65-year-old, respectively (p-value <10^−3^, Table 2.). We estimated R_0_ to 10.6 (95%CI: 8.4-12.7, Table 2.). The viral production p was estimated to 1.20×10^6^ viruses per day (95%CI: 6.5×10^5^-1.7× 10^6^), leading to a burst size of infectious viruses, given by 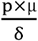 of 1.36×10^2^ and 1.60×10^2^ in individuals aged < 65-year-old and those aged ≥ 65-year-old, respectively.

**Table 2.** Parameters distribution using model averaging (Median, 95%CI). *R*_0_: basic reproductive number; *δ*: loss rate of infected cells; *p*: rate of viral production; □: remdesivir effect

| Parameter estimates |  |  |
| --- | --- | --- |
| Parameter | Fixed effects<br>(Median, 95% CI) | SD of the random effect<br>(Median, 95% CI) |
| $R_0$ | 10.60 (8.53-12.68) | 0.50 |
| $\delta_{<65}$ ($d^{-1}$) | 0.88 (0.80-0.96) | 0.46 (0.41-0.51) |
| $\delta_{\geq 65}$ ($d^{-1}$) | 0.76 (0.67-0.84) | |
| $p$ ($10^6$ virus.cell$^{-1}$.d$^{-1}$)* | 1.20 (0.66-1.72) | 0.38 (0.14-0.63) |
| $\epsilon$ (%) | 52 (35-69) | 0.77 (0.18-1.37) |
| $\sigma$ (log$_{10}$ RNA copies/$10^4$ cells) | 1.14 (1.09-1.19) | - |

\* the value of $p$ depends on the normalization factor used to quantify viral load (see methods)

### Estimation of remdesivir effect on viral dynamics

Remdesivir effect, ε, was estimated for different putative values of the pharmacological delay ranging from 0 to 5 days (see methods). Using model averaging, we estimated remdesivir to decrease the viral production p by ε =52%, (95%CI: 35-68%, Table 2.), i.e., a 2-fold reduction compared to SoC, and this effect was statistically significant in model averaging (p-value = 0.0031). Of note, the antiviral effect of remdesivir was statistically significant for τ = 0 to τ = 3, with values equal to 49%, 50%, 53% & 52%, respectively (p-value = 0.020, 0.033, 0.0026, 0.0024 respectively, Fig. 3., Supplementary Table S1.).

**Fig. 2.**
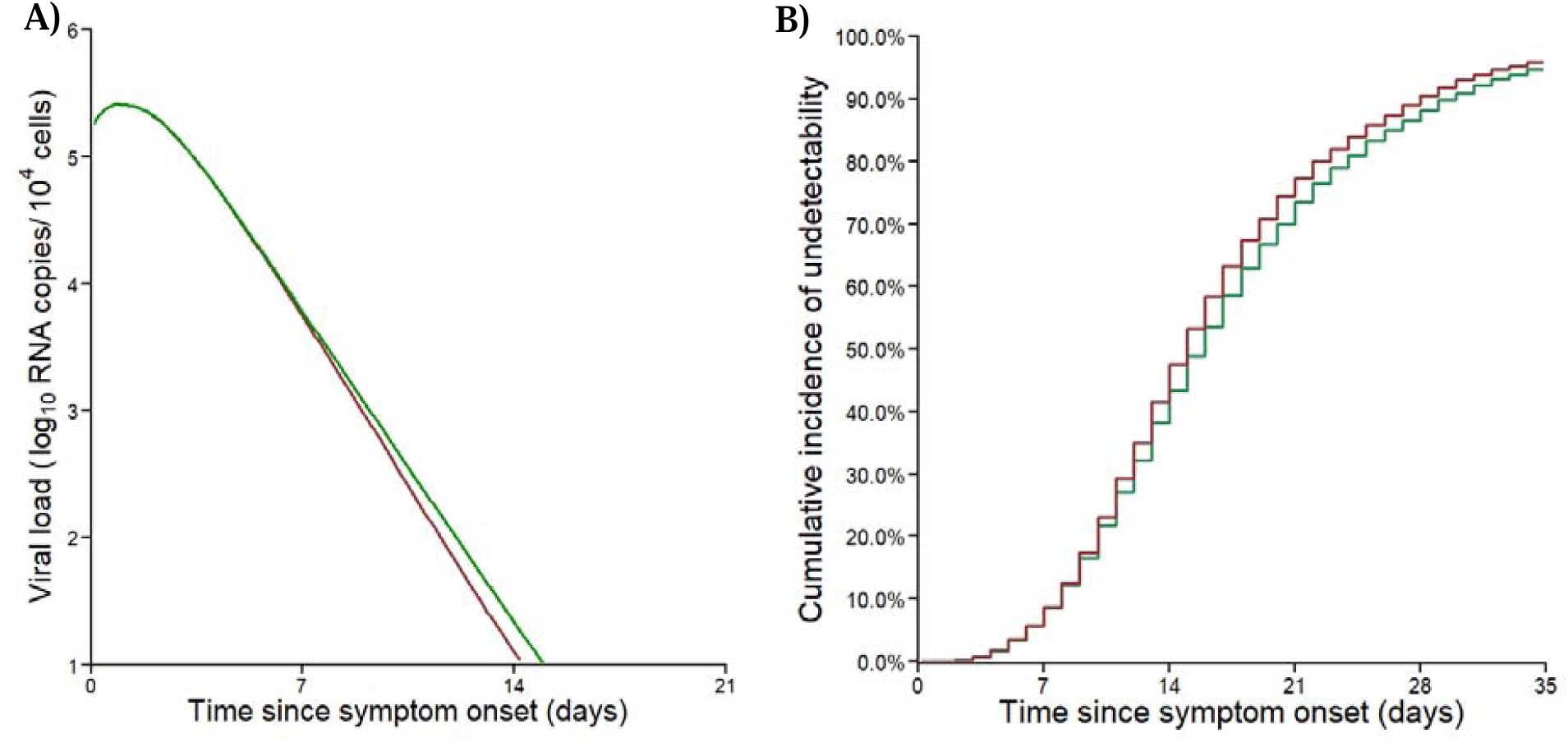
Viral dynamics predicted by the model. A) Median predicted nasopharyngeal viral dynamics according to the time since symptom onset. B) Cumulative incidence of the predicted time to viral clearance. Red: patients receiving remdesivir + SoC. Green: patients receiving SoC only. Results obtained by sampling 5,000 individuals in the estimated parameter distributions.

**Fig. 3.**
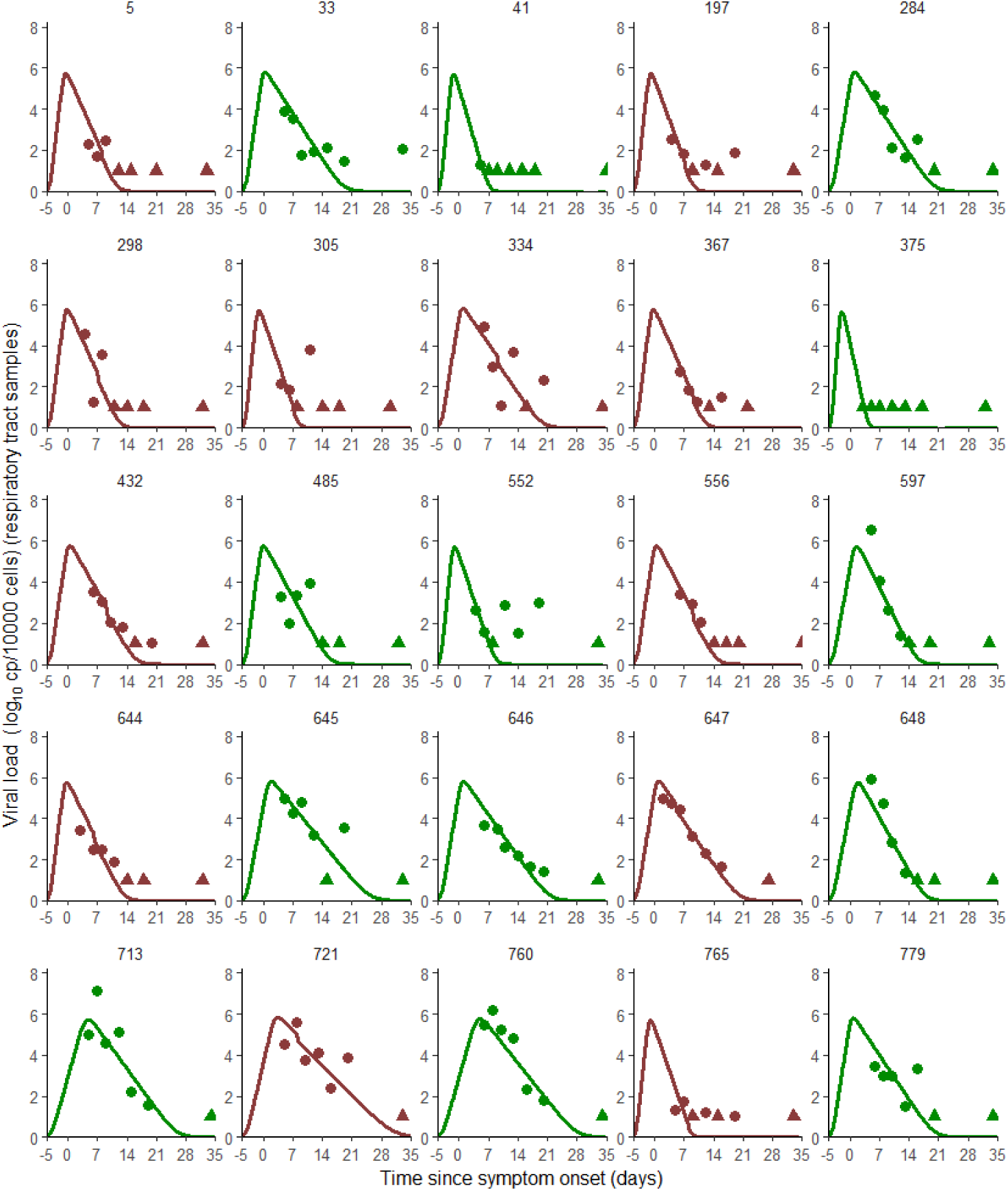
Model-based individual fits for τ= 3. Patients represented here are the patients with 7 data points admitted within the first week of symptom onset. Circles represent detectable viral load, triangles represent data below the limit of quantification. Results obtained by using the individual predictions of the model τ=3. Red: patients receiving remdesivir + SoC. Green: patients receiving SoC only.

To get a sense of the effect size, we simulated 5,000 *in silico* virological profiles using the parameters distributions estimated by the model and we calculated the exact time to viral clearance for each simulated patient. Remdesivir administration shortened the time to viral clearance by 0.7 day (IQR:0.0-1.3, with a median time to viral clearance equal to 14.5 days (IQR: 10.4-20.2 days) and 15.3 days since symptom onset (IQR: 10.6-21.5 days) in treated and untreated individuals, respectively (Fig. 2a-b.). Patients older than 65-year-old, having a diminished loss rate of infected cells (p-value<10^−3^) consequently had a longer median time to viral clearance than those <65, both in treated and untreated patients (15.6 vs 16.6 days in treated and untreated individuals, respectively, for patients ≥ 65-year-old and 13.3 vs 14.1 days in treated and untreated individuals, respectively, for patients < 65-year-old). In terms of time since randomization, remdesivir led to a time of viral clearance also reduced by 1 day, with a median time equal to 5.5 days (IQR: 1.0-12.0 days) and 6.5 days (IQR: 1.0-13.0 days) in treated and untreated individuals, respectively (Supplementary Fig. S3a&c.). Overall, the maximal difference in viral load between treated and untreated individual was obtained at day 11.5 post symptom onset, with a difference in median viral loads of 0.24 log_10_ copies/10^4^ cells (IQR: 0.06-0.39), corresponding to a 2-fold reduction in viral load levels (Fig. 2a.).

We repeated the same analysis in subpopulations stratified by their time of treatment initiation ≤ or > 7 days since symptom onset. In patients in whom treatment was initiated in the first 7 days following symptom onset (N=210, Supplementary Fig. S4.), remdesivir effect was statistically significant for τ=3 (ε=36%; p-value=0.041, Supplementary table S2a.), but was not significant when averaging the different pharmacological delays (p-value =0.07, Supplementary table S2b.). In patients arriving >7 days after symptom onset and later (N=455), remdesivir efficacy ε was not significant in any model M_i_ (Supplementary table S3a.), and consequently in model averaging (Supplementary table S3b.).

In patients with a high viral load at admission (See Methods, N=184, median time of admission = 8 days since symptom onset, IQR: 6-10, Fig. 1b.), remdesivir was estimated to reduce viral production by 80% (95%CI: 65-96%), i.e., a reduction by 5-fold compared to SoC, and this effect was statistically significant in model averaging (p-value < 10^−5^, see Methods, Supplementary table S4a.). In line with the analysis on the whole population, remdesivir effect was statistically significant for values of τ ranging from 0 to 3 days after treatment initiation, with values equal to 80%, 82%, 50% and 55% respectively (p-value <10^−3^ for all values of τ, Supplementary table S4b.). In patients admitted with a low viral load (N=236, median time of admission = 9 days since symptom onset, IQR: 7-12, Fig. 1c.), remdesivir effect was lower and was estimated to 49%, with a poor precision of the estimation (95%CI: 0-100, p-value =0.013, Supplementary table S5a.).

Using parameters distributions estimated in the population with high viral load at randomization, simulations showed that remdesivir led to a time to viral clearance shortened by 2.4 days (IQR:0.9-4.5, Fig. 4.). Further, remdesivir led to a maximal difference between median viral loads of treated and untreated patients of 0.7 log_10_ copies/10^4^ cells, reached at 12 days after symptom onset. Similar effects were obtained when looking at data in terms of time since randomization, with a median time equal to 10.5 days (IQR: 6.5-14.6 days) and 13.5 days (IQR: 10.0-18.0 days) in treated and untreated individuals, respectively (Supplementary Fig. S3b&d.).

**Fig. 4.**
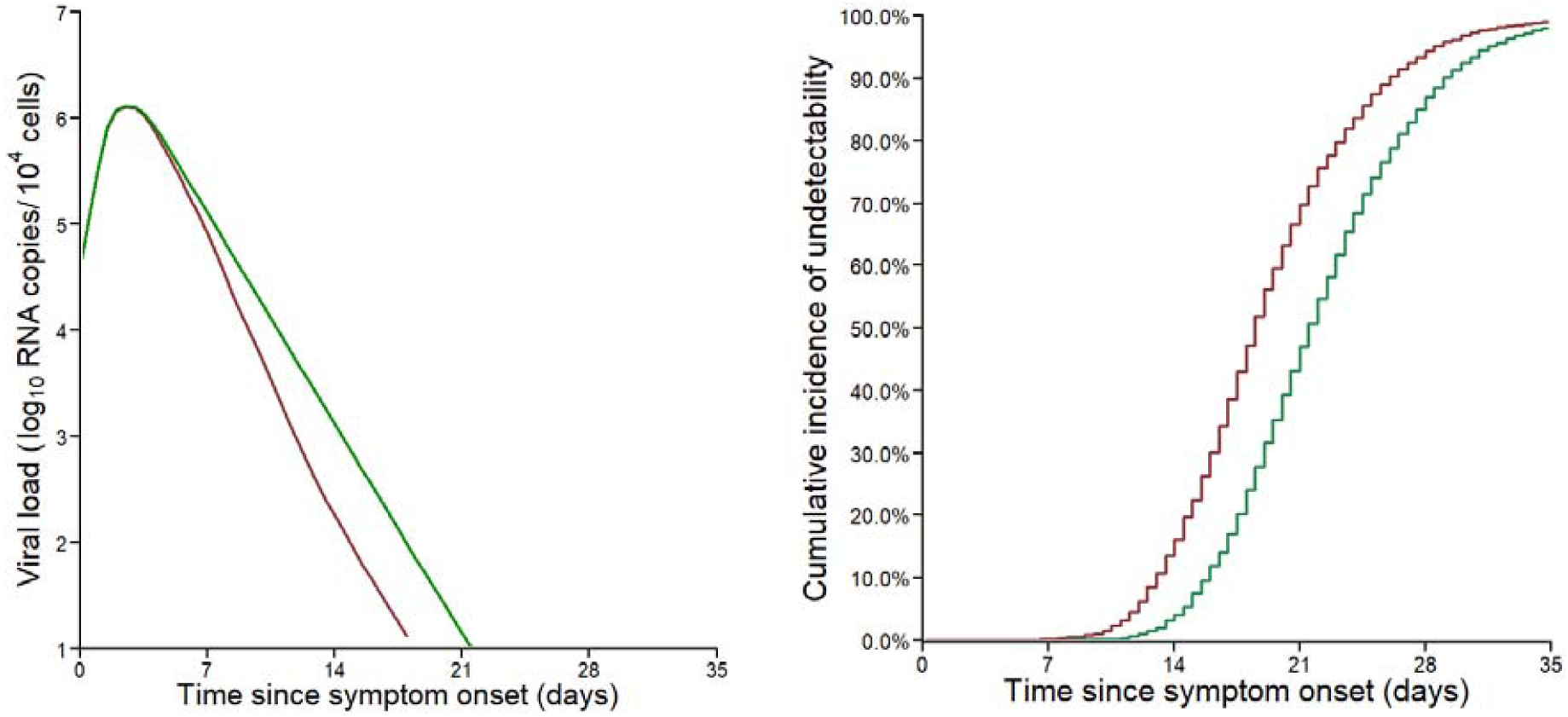
Viral dynamics predicted by the model in subpopulation of viral load at admission 3.5 log_10_ copies/10^4^ cells. A) Median predicted nasopharyngeal viral dynamics according to the time since symptom onset. B) Cumulative incidence of the predicted time to viral clearance. Red: patients receiving remdesivir + SoC. Green: patients receiving SoC only. Results obtained by sampling 5,000 individuals in the estimated parameter distributions.

## Discussion

We found that remdesivir had a statistically significant antiviral efficacy, reducing viral production from infected cells by 52% (95%CI: 35-68%), corresponding to a 2-fold reduction in viral production. Despite this, the result is a modest impact on viral dynamics, with an average 0.7 day reduction in the time to viral clearance and large interindividual variability. It explains why less sophisticated approaches did not find any effect of remdesivir on viral dynamics *(6, 9, 19)*.

The overall limited antiviral effect of remdesivir may be due to factors that mitigate a potential effect, in particular the heterogeneity in the time between infection and treatment initiation as well as the variability in viral load levels at the time of treatment initiation. To address this question, we here relied on a very rich dataset of 665 patients, for which viral loads were centralized and normalized, and we used a model previously developed on a large cohort of hospitalized patients to disentangle the effects of remdesivir from the natural clearance of viral load. To identify factors associated with the effects of remdesivir on viral kinetics, we performed exploratory analyses by stratifying the population on timing of treatment initiation in days since symptom onset and on viral load at randomization, the latter being an exploratory analysis not included in the initial trial protocol. We did not identify a significant effect of remdesivir in patients treated early, consistent with the results found earlier, using descriptive models *(19)*. However, in the subset of patients in which viral load at admission was higher than 3.5 log_10_ copies/10^4^ cells, remdesivir effect in reducing viral production was greater than in the overall population (80% vs 52%, respectively), leading to a median reduction of 2.4 days (IQR:0.9-4.5) in the time to viral clearance. These results therefore suggest that remdesivir could be more effective in patients with high viral load, at a stage of the infection where viral replication is very active. The larger antiviral effect observed in patients with higher viral loads is also consistent with the results of the Recovery trial *(4)* that identified a significant effect of monoclonal antibody combination REGN-CoV-2 on clinical outcome in hospitalized patients with a seronegative status at baseline, a factor associated with high viral load *(2, 20)*. The relationship between treatment effect and viral load was also reported in outpatients receiving monoclonal antibodies within 7 days of symptom onset *(2, 20)*. Simulations conducted with our model suggested no dramatic difference in viral shedding when administration is initiated between 3-7 days (Supplementary Fig. S5.), but our model did not account for immune mechanisms that could act, in concert with early antiviral treatment, to accelerate viral clearance *(21)*. Taken together, these data still suggest that it could be beneficial to administrate remdesivir as early as possible. Consistent with this interpretation, preliminary results from the Pinetree study showed that administration of remdesivir in outpatients within 7 days of symptom onset led to a significant reduction of the risk of hospitalization *(22)*. However, it should be acknowledged that preliminary data did not find any effect of remdesivir on viral kinetics.

How does remdesivir antiviral efficacy compare with other antiviral drugs now available? Our estimated effect of 52% in the global population is below the pharmacodynamic target of 90% that we determined previously to achieve a clinical efficacy in hospitalized patients and/or to prevent disease acquisition in prophylactic setting *(15, 17, 23)*. It is also probably lower than what has been shown in outpatients treated with monoclonal antibodies. For instance, 7 days after treatment initiation, the viral load in ambulatory individuals receiving casirivimab+imdevimab or bamlanivimab+etesevimab was reduced by 5-10 fold as compared to patients receiving placebo, as compared to 2 fold in our hospitalized population here *(2, 24)*. Using a viral dynamic model, we estimated that the effect of COVA1-18, another monoclonal antibody, was greater than 95% in blocking viral infection in a macaque model SARS-CoV-2 infection *(25)*. The lower efficacy of remdesivir could nonetheless be relevant in a combination setting to increase the genetic barrier to resistance due to the emergence of variants of concerns, and the overall efficacy as suggested in the mice model *(26)*. This could be even more relevant in patients with high viral load to reduce the risk of mutation emergence in a context where huge quantities of viral progeny are still produced upon treatment initiation.

We acknowledge some important limitations to our study. First a more complete evaluation of remdesivir would involve the analysis of viral dynamics in the lower respiratory tract, as was done in non-human primates *(12, 13)*. Here, viral loads in lower respiratory tracts were available in a subset of 120 individuals. However, the number of samples were limited and these individuals had a very severe disease (Supplementary table S6.), making it not possible to provide an unbiased and precise estimate of remdesivir. Second, we could not evaluate the association between remdesivir drug concentration and viral decay, which would be important to draw more definitive evidence on remdesivir antiviral activity. Here, drug concentrations were available for only a limited number of patients (N=61), and no significant association between drug concentrations and the time to viral clearance (Supplementary Fig. S6.) could be found. Moreover, given that symptom onset are posterior to the peak viral load *(17, 27)*, a potential bias in the estimation of early viral dynamic parameters cannot be ruled out (see a discussion on that aspect in Néant et al *(17)*), in particular in hospitalized patients. Finally, the use of adjuvant drugs such as corticosteroids or any immunosuppressive treatment which might promote viral replication, as well as the intrinsic immune competency of each treated individual have not been considered.

In conclusion, the use of a within-host model of the infection allowed us to estimate the *in vivo* antiviral efficacy of remdesivir in the nasopharyngeal swabs of hospitalized patients. Overall, we showed that remdesivir had a modest antiviral activity in reducing viral production, leading to a reduction by 0.7-day of the time to viral clearance (IQR:0.0-1.3). In exploratory analyses, remdesivir had a higher antiviral efficacy in patients with high viral load at randomization (≥3.5 log copies/ 10^4^ cells), leading to a reduction of time to viral clearance of 2.4 (IQR:0.9-4.5) days between treated and untreated patients. The limited effect of remdesivir is consistent with the lack of clinical efficacy in hospitalized patients reported by the DisCoVeRy trial *(19)*.

### Patients and methods

#### Study design and data collection

Hospitalized patients with a laboratory-confirmed SARS-CoV-2 infection were enrolled in the DisCoVeRy trial (NCT 04315948; EudraCT 2020-000936-23, *(19)*), sponsored by the Institut national de la santé et de la recherche médicale (Inserm, France). We analyzed the results obtained from patients allocated to receive either SoC alone or in addition to remdesivir between March 2020 and January 2021, hospitalized in 48 different sites from France, Belgium, Portugal, Austria and +Luxembourg, and for whom nasopharyngeal swabs were available. Patients who were randomized more than 20 days after symptoms onset were removed from analysis to limit bias in the estimation of the viral dynamic parameters. Remdesivir was administrated intravenously at a loading dose of 200 mg on day 1 followed by 100 mg infusions once-daily for up to 10 days. More details on the study design, ethic approval and inclusion/exclusion criteria can be found in *(19)*.

#### Viral load measurements

Normalized viral load in samples were measured at randomization and at days 3, 5, 8, 11, 15±2 and 29±3 after randomization, in nasopharyngeal swabs collected through validated devices containing flocked swabs and virus transport medium. Viral load was also assessed in broncho-alveolar lavage (BAL) fluids for a subset of patients, upon clinician’s discretion. Viral and cellular loads were all centralized in the National Centre for Viral Respiratory Infections (Hospices Civils de Lyon, France) and determined by real time quantitative reverse transcriptase polymerase chain reaction (RT-PCR) blinded to treatment arm. To allow the comparison of samples with different number of cells and from different sampling sites (nasopharyngeal swabs and lower respiratory samples), we used a normalized SARS-CoV-2 viral load by dividing the viral load measured in the sample by the number of cells measured and expressed it copies per 10^4^ cells. We estimated the limit of detection (LoD) to 1 log_10_ copies/10^4^ cells and all viral loads strictly below LoD were considered as censored. More details on viral load quantification techniques can be found in *(19)*.

### Viral dynamic model

#### Model equations

Given the small number of patients hospitalized in the first days after symptom onset *(19)*, we used a simple target-cell limited model with an eclipse phase to characterize viral dynamics *(17)*, and did not consider more complex models including an immune response *(17)*. The model includes target cells (T) and infected cells, that are initially non-productive (I_l_) during the eclipse phase and become subsequently productively infected (I_2_). Such a model assumes that target cells are infected at a constant infection rate β, and the mean duration of the eclipse phase is noted 1/k. We assumed that productively infected cells had a constant loss rate, noted δ, that differs between patients above or below 65 years old *(17)*. Each infected cell produces p viral particles per day, but only a fraction of these particles, µ, is infectious. We note V the sum of the infectious viral particles, noted V_i_, and of the non-infectious viral particles, noted V_ni_, and both are cleared at a rate c. The model can be written as:

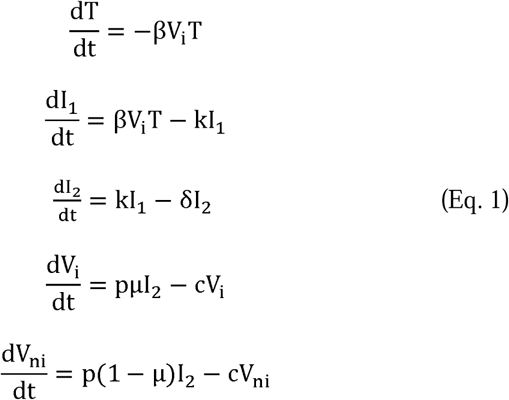

At the time of infection, we assumed that there is exactly one productively infected cell I_2_ in the entire nasopharyngeal tract, thus T=T_0_; I_1_=0; I_2_=1; V_i_=0; V_ni_=0. The basic reproductive number R_0_, defined as the number of secondary infected cells resulting from one infected cell in a population of fully susceptible cells, is equal to βpT_0_µ/cδ, where T_0_, the total number of susceptible cells, is equal to 4 × 10^6^ cells *(28)*. In the same manner, one can derive the infectious burst size, 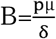. We used a scaling factor, f, to convert V into a normalized viral load expressed in copies/10^4^ cells, and we note V_obs_= f×V. As only the parameter p×f can be estimated, we assumed without loss of generality that the proportion of susceptible cells in the biological sample was on average 10 fold lower than in the nasopharyngeal compartment, i.e., 0.1% *(28)*. Thus, we fixed f to 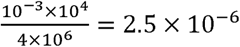. Changing the value of the scaling factor would affect the estimate of p, but not the value of the other parameters, including the estimation of R_0_ and the treatment efficacy (see below).

We fixed c to 10 d^-1^, k to 4 d^-1^ and µ to 10^−4^ as previously published *(15, 17, 21)*. Further we fixed the time of infection to 5 days prior to symptom onset in all individuals, which corresponds to the mean duration of the incubation phase *(17)*. We estimated p and δ as well as their inter individual variabilities. All viral dynamic parameters were assumed to follow a lognormal distribution. Given the lack of data in the viral upslope, we also fixed the inter individual variability of R_0_ to 50%.

#### Modeling the effect of remdesivir and pharmacological delay

The initiation of remdesivir reduces viral production p by a factor ε, a parameter following a logistic distribution to ensure values comprised between 0 (no activity) and 1 (full viral suppression), leading to:

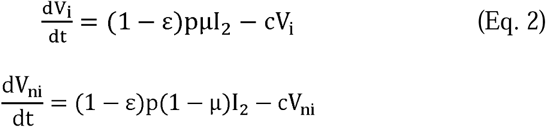

Active forms of remdesivir have a long mean terminal elimination half-life (t_1/2_) of about 24h, and in non-human primates reaches steady state concentrations at day 4 *(29)*. Therefore, we aimed to incorporate a potential delay between remdesivir administration and its effect, and assumed that treatment was only effective after a certain delay after treatment initiation τ. We considered different possible values for τ, ranging from τ=0 (model M_0_) to τ=5 (model M_5_).

We used a model averaging approach *(30)* to account for the uncertainty on τ. Thus, the median parameter estimates and their 95% confidence intervals were obtained by sampling in the mixture of the parameter estimate asymptotic distribution of each model, with weights of model M_i_ calculated as follows: 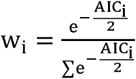 where AIC_i_ is the Akaike Information Criterion (AIC) of model M_i_. We computed the model averaged likelihood using weights M_i_ and tested the significance of the effect of remdesivir in both models M_i_ and model averaging using Likelihood Ratio Test (LRT) *(31)*.

To quantify the effect of remdesivir on viral clearance (i.e., when the predicted viral load is lower than 1 log_10_ copies/10^4^ cells), we simulated 5,000 treated individual virologic profiles in the parameter asymptotic distribution of the parameters with each model M_i_ associated with probability w_i_. Times of treatment initiation were sampled from the fitted Gamma distribution of the observed times of randomization in the population. For each simulated individual, we then calculated the area under curve (AUC) of viral load and the time to reach viral clearance, and we computed the cumulated incidence of viral clearance. The same individual parameters were used assuming a treatment efficacy, ε, equal to 0, to simulate 5000 untreated individuals and compute the difference between a treated individual and its own control. Finally, we evaluated the effects of earlier treatment initiation, assuming a fixed time to treatment initiation of 3, 5, 7, and 9 days after symptom onset.

#### Sub-analyses according to viral load at admission and time since symptom onset

Because the timing of treatment initiation is a key factor for antiviral drug efficacy *(15)*, we used the same modeling strategy in two distinct populations, e.g, in patients in whom treatment was initiated early (≤7 days since symptom onset) and those initiating the treatment (>7 days since symptom onset). We chose the cutoff of 7 days from symptom onset, consistent with *(19)*. Similarly, we conducted the same analysis in patients with high or low viral load at admission, taking the cutoff of 3.5 log_10_ copies/10^4^ cells. This value represents the threshold level below which the virus cannot be cultured and is unlikely infectious *(32)*.

#### Parameter estimation & fitting assessment

Parameters were estimated by maximum likelihood estimation using the stochastic approximation expectation–maximization (SAEM) algorithm implemented in Monolix Software 2020R1 (http://www.lixoft.eu).

## Data Availability

All data produced in the present study are available upon reasonable request to the authors

## Supplementary Materials

### Supplementary Materials and Methods

Plasma drug determination

### Supplementary figures and tables

**Fig. S1**. Flowchart of data selection in viral kinetic modeling

**Fig. S2**. Distribution of randomization times since symptom onset

**Fig. S3**. Viral dynamics predicted by the model in time since randomization

**Fig. S4**. Nasopharyngeal viral load data in 665 patients from DisCoVeRy trial analyzed in the present study.

**Fig. S5**. AUC of viral loads for 5000 simulated patients treated 3, 5, 7 or 9 days after symptom onset, or left untreated.

**Fig. S6**. Pharmacokinetic/pharmacodynamic effect.

**Table S1**. Population parameters of models including remdesivir efficacy starting between 0 and 5 days after randomization.

**Table S2a**. Population parameters of models including remdesivir efficacy starting between 0 and 5 days after randomization in patients randomized ≤7 days since symptom onset.

**Table S2b**. Parameters distribution using model averaging (Median, 95% CI) in patients randomized ≤7 days since symptom onset.

**Table S3a**. Population parameters of models including remdesivir efficacy starting between 0 and 5 days after randomization, in patients randomized >7 days since symptom onset.

**Table S3b**. Parameters distribution using model averaging (Median, 95% CI) in patients randomized>7 days since symptom onset.

**Table S4a**. Parameters distribution using model averaging (Median, 95% CI) in patients with viral load at admission ≥3.5 log_10_ copies/10^4^ cells.

**Table S4b**. Population parameters of models including remdesivir efficacy starting between 0 and 5 days after randomization in patients with viral load at admission ≥3.5 log_10_ copies/10^4^ cells.

**Table S5a**. Parameters distribution using model averaging (Median, 95% CI) in patients with viral load at admission < 3.5 log_10_ copies/10^4^ cells.

**Table S5b**. Population parameters of models including remdesivir efficacy starting between 0 and 5 days after randomization, in patients with viral load at admission < 3.5 log_10_ copies/10^4^ cells.

**Table S6**. Characteristics of the population with in lower respiratory tract samples.

## Fundings

The trial is funded by grants from:

European Commission (EU-Response, Grant 101015736)

Programme Hospitalier de Recherche Clinique (PHRC-20-0351, Ministry of Health)

Domaine d’intérêt majeur One Health Île-de-France (R20117HD)

REACTing, a French multi-disciplinary collaborative network working on emerging infectious diseases.

Fonds Erasme-COVID-Université Libre de Bruxelles

Belgian Health Care Knowledge Centre

Austrian Group Medical Tumor

European Regional Development Fund

Portugal Ministry of Health

Portugal Agency for Clinical Research and Biomedical Innovation.

## Author contribution

Methodology: AG, GP, MBD

Investigation: MH, TS, RG, JAP, JP, NPS, DC, YY, FW, AGB, FM, FA, CB

Project administration: JG

Supervision: JG, MBD

Writing – original draft: GL, NN

Writing – review & editing: AG, DB, NPS, DC, FW, AGB, FM, FA, CB, JG, MBD

## Data and materials availability

All data and code will be available upon publication

