## Supplementary material for "Modeling remdesivir antiviral efficacy in COVID-19 hospitalized patients of the randomized, controlled, open-label DisCoVeRy trial"

^1^Université de Paris, IAME, INSERM, France ; ^2^ Laboratoire de Virologie, Institut des Agents Infectieux de Lyon, Centre National de Référence des virus respiratoires France Sud, Hospices Civils de Lyon, F-69317, Lyon, France; ^3^Université de Lyon, Virpath, CIRI, INSERM U1111, CNRS UMR5308, ENS Lyon, Université Claude Bernard Lyon 1, F-69372, Lyon, France ; ^4^AP-HP, Hôpital Bichat, Département d'Épidémiologie, Biostatistique et Recherche Clinique, F-75018, Paris, France; CIC-EC 1425, INSERM, F-75018, Paris, France ; ^5^AP-HP, Hôpital Bichat Claude Bernard, Laboratoire de Pharmacologie-toxicologie, F-75018 Paris, France ; ^6^Cliniques Universitaires de Bruxelles – Hôpital Érasme, Université Libre de Bruxelles, Clinique des maladies infectieuses, Brussels, Belgium ; ^7^Centre hospitalier de Luxembourg, Service des maladies infectieuses, L-1210 Luxembourg, Luxembourg ; ^8^Department of Internal Medicine III with Haematology, Medical Oncology, aemostaseology, Infectiology and Rheumatology, Oncologic Center, Salzburg Cancer Research Institute - Laboratory for Immunological and Molecular Cancer Research (SCRI-LIMCR), Paracelsus Medical University Salzburg, 5020 Salzburg, Austria ; ^9^Cancer Cluster Salzburg, 5020, Salzburg, Austria ; ^10^AGMT, 5020 Salzburg, Austria. ; ^11^Centro Hospitalar São João, Emergency and Intensive Care Department, Porto, Portugal ; ^12^Universidade do Porto, Faculty of Medicine, Porto, Portugal ; ^13^Université de Lille, Inserm U1285, CHU Lille, Pôle de réanimation, CNRS, UMR 8576 - UGSF - Unité de Glycobiologie Structurale et Fonctionnelle, F-59000, Lille, France ; ^14^AP-HP, Hôpital Bichat, Service de Maladies Infectieuses et Tropicales, F-75018 Paris, France ; ^15^National Institute for Health Research, Health Protection Research Unit in Healthcare Associated Infections and Antimicrobial Resistance, Imperial College London, London, UK ; ^16^Sorbonne Université, Inserm, Institut Pierre-Louis d'Épidémiologie et de Santé Publique, F-75013, Paris, France ; ^17^Hospices Civils de Lyon, Hôpital Lyon-Sud Pierre-Bénite, Département de Soins Intensifs, F-69000, Lyon, France ; ^18^CHU de Saint-Etienne, Service d’Infectiologie, F- 42055 Saint-Etienne, France ; ^19^Université Jean Monnet, Université Claude Bernard Lyon 1, GIMAP, CIRI, INSERM U1111, CNRS UMR5308, ENS Lyon, F-42023 Saint-Etienne, France ; ^20^CIC 1408, INSERM, F- 42055 Saint-Etienne, France ; ^21^Hospices Civils de Lyon, Département des maladies infectieuses et tropicales, F-69004, Lyon, France ; ^21^Université Claude Bernard Lyon 1, CIRI, INSERM U1111, CNRS UMR5308, ENS Lyon, F-69372, Lyon, France

† equally contributed

**Keywords:** *SARS-CoV-2; Viral dynamics; remdesivir; modeling*

**This PDF file includes:**

**Supplementary Materials and Methods**

**Supplementary figures S1 to S4**

**Supplementary tables S1 to S6**

**Materials and Methods:**

***Plasma drug determination***

Remdesivir requires triphosphorylation by intracellular kinases to be pharmacologically active. After administration, it is metabolized into two main sub-compounds: first GS-441524, the predominant active plasma metabolite, and then GS-443902, which is the PBMC-associated pharmacologically most active metabolite. In a subset of patients from several centers, plasma samples were collected and centralized to measure remdesivir and GS-441524 at day 1 for maximal concentration (C_max_) and at days 2, 5 and 8 after treatment initiation for trough concentrations (C_trough_). Both remdesivir and GS-441524 were measured using Liquid Chromatography coupled with tandem Mass Spectrometry, after plasma protein’s precipitation with a lower limit of quantification at 1 ng/mL.

### Fig. S1. Flowchart of data selection in viral kinetic modeling

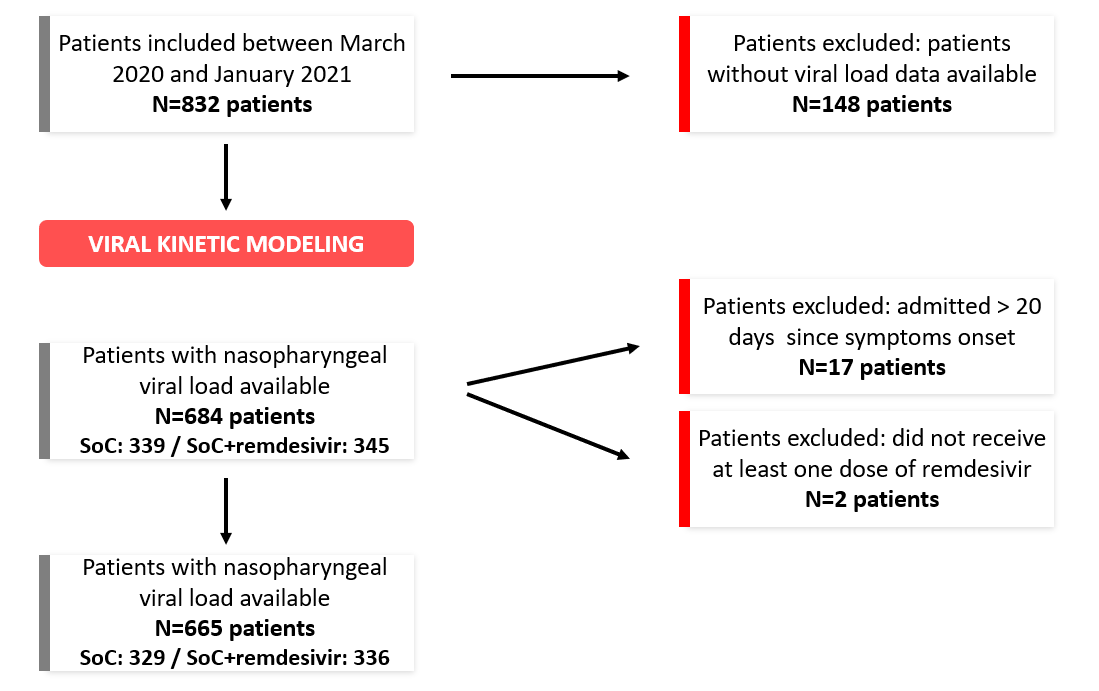

### Fig. S2. Distribution of randomization times since symptom onset. Red: patients receiving remdesivir + SoC. Green: patients receiving SoC only.

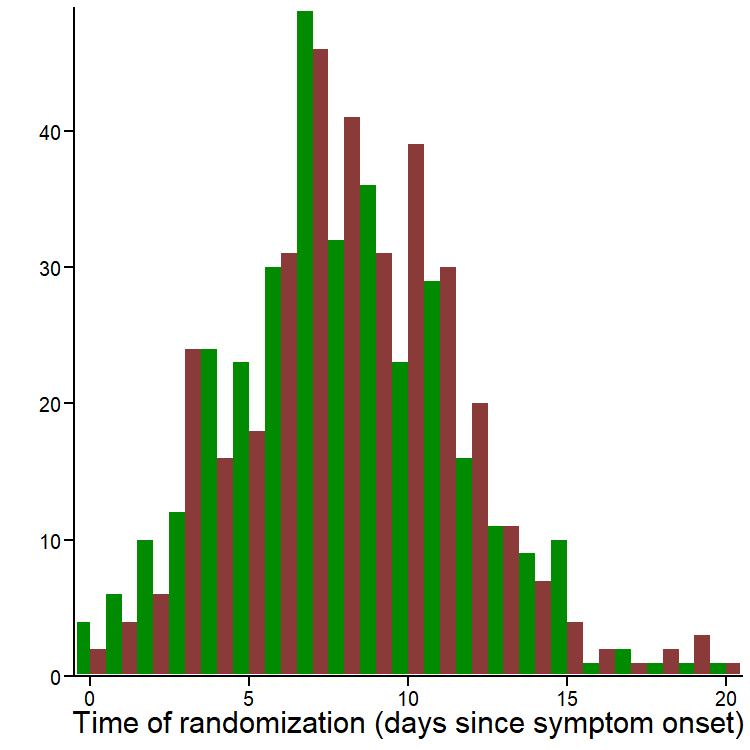

**Fig. S3. Viral dynamics predicted by the model in time since randomization**. Top: Median predicted nasopharyngeal viral dynamics according to the time since randomization. Bottom: Cumulative incidence of the predicted time to viral clearance. Left: Whole population. Right: Patients with viral load at baseline $\geq$ 3.5 log_10_ copies/10^4^ cells. Red: patients receiving remdesivir + SoC. Green: patients receiving SoC only. The simulated individuals presented here are the same as in Figure 2 & 4, centered on time of randomization.

b
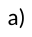
)

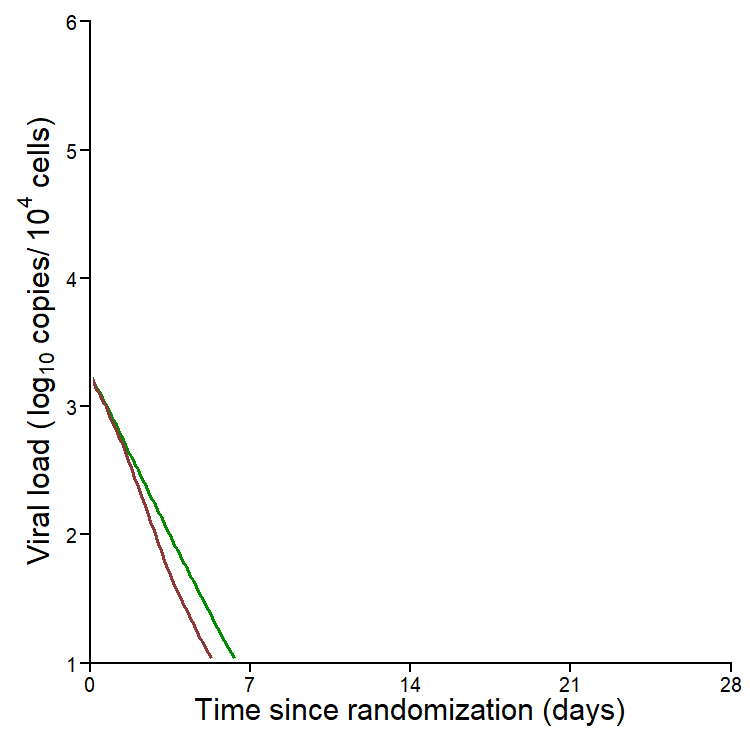

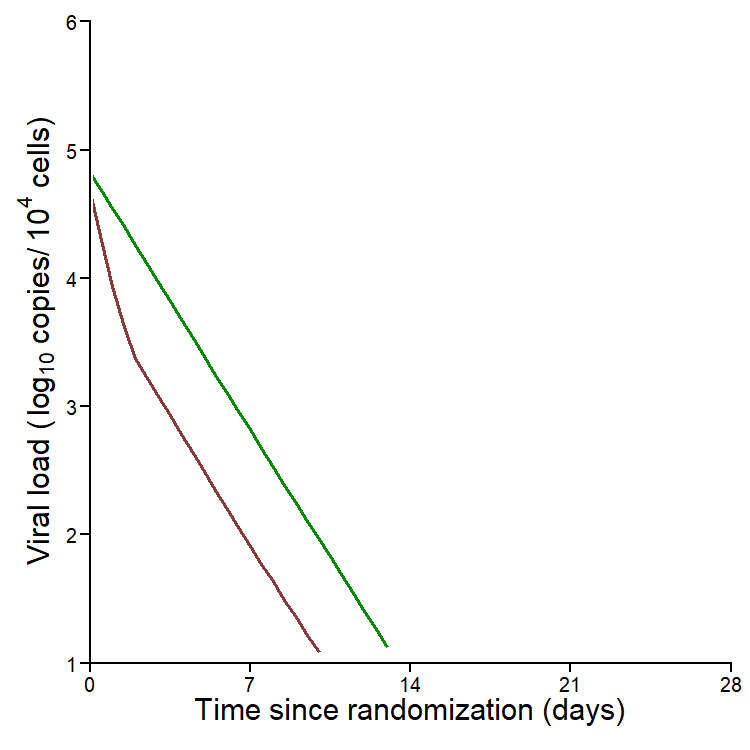

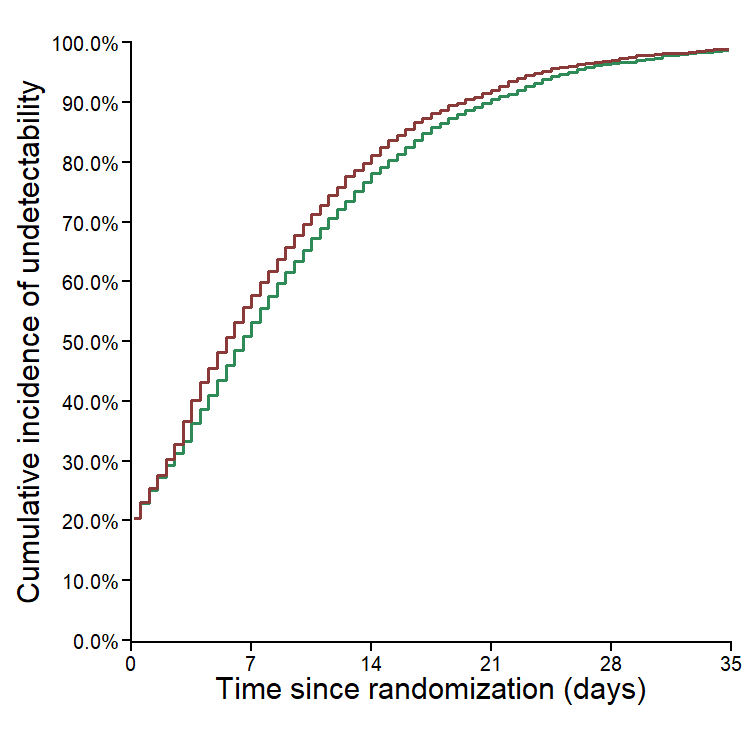

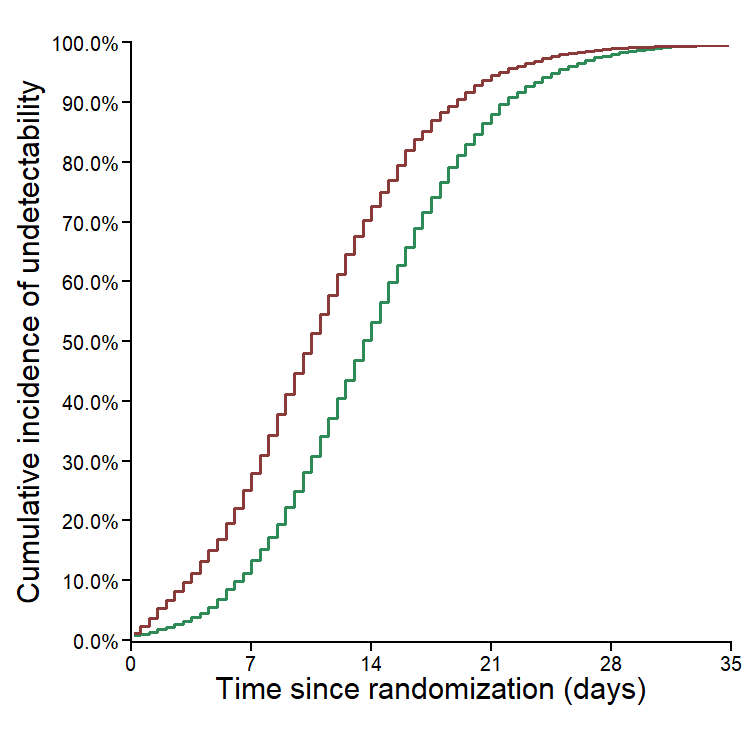

a)

c)

d)

**Fig. S4. Nasopharyngeal viral load data in 665 patients from DisCoVeRy trial analyzed in the present study.** SARS-CoV-2 nasopharyngeal viral load according to the time since randomization in patients admitted within the first week of symptom onset (N=210, a) and after the first week of symptom onset (N=455, b). Data are presented as means (95%CI). Red: patients receiving Remdesivir + SoC. Green: patients receiving SoC only.

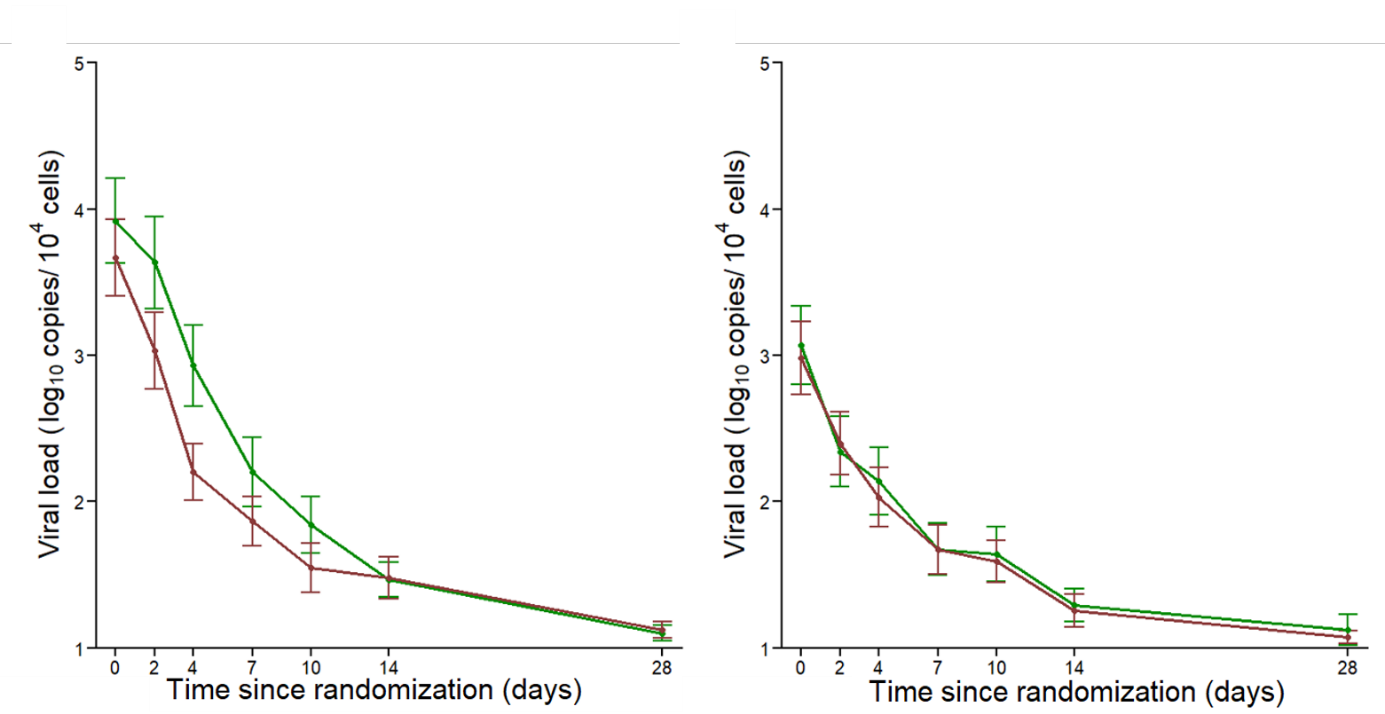

b)

a)

### Fig. S5. AUC of viral loads for 5000 simulated patients treated 3, 5, 7 or 9 days after symptom onset, or left untreated. a) Whole population. b) Subpopulation with viral load at baseline $\boldsymbol{\geq}$ 3.5 log_10_ copies/10^4^ cells. c) Subpopulation with viral load at baseline < 3.5 log_10_ copies/10^4^ cells. The individual parameters are identical to the ones used in Figure 2 & 4, except for the time of treatment initiation.

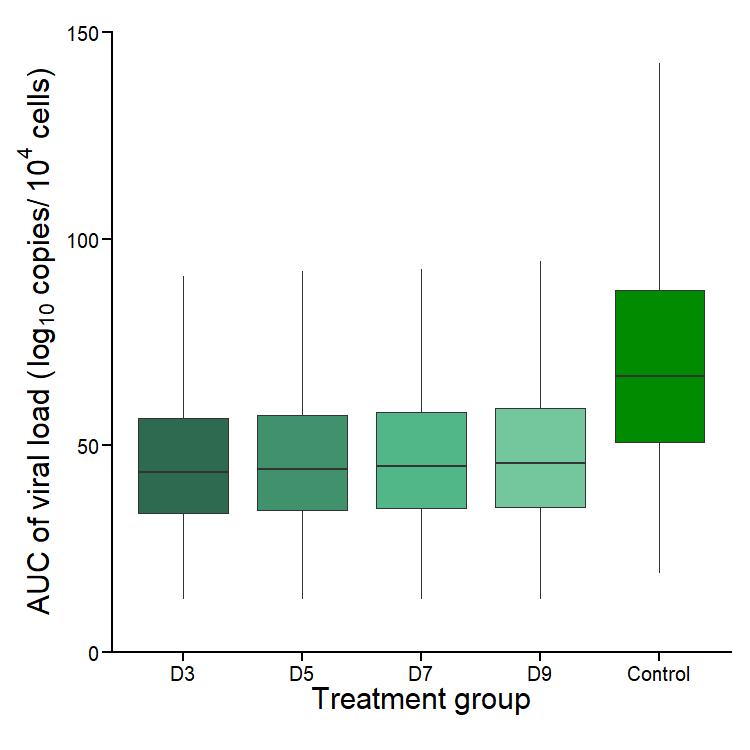

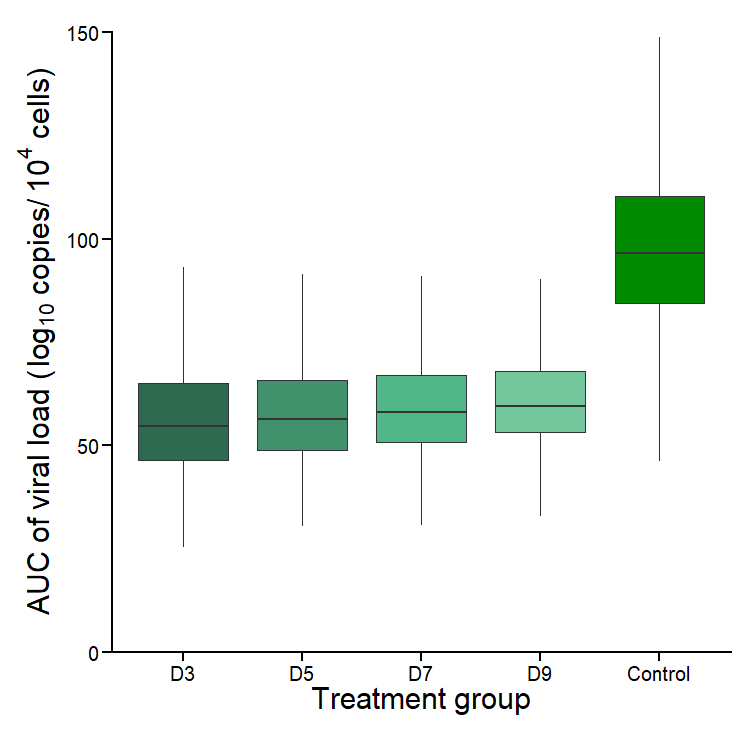

b)

a)

### Fig. S6. Pharmacokinetic/pharmacodynamic effect.

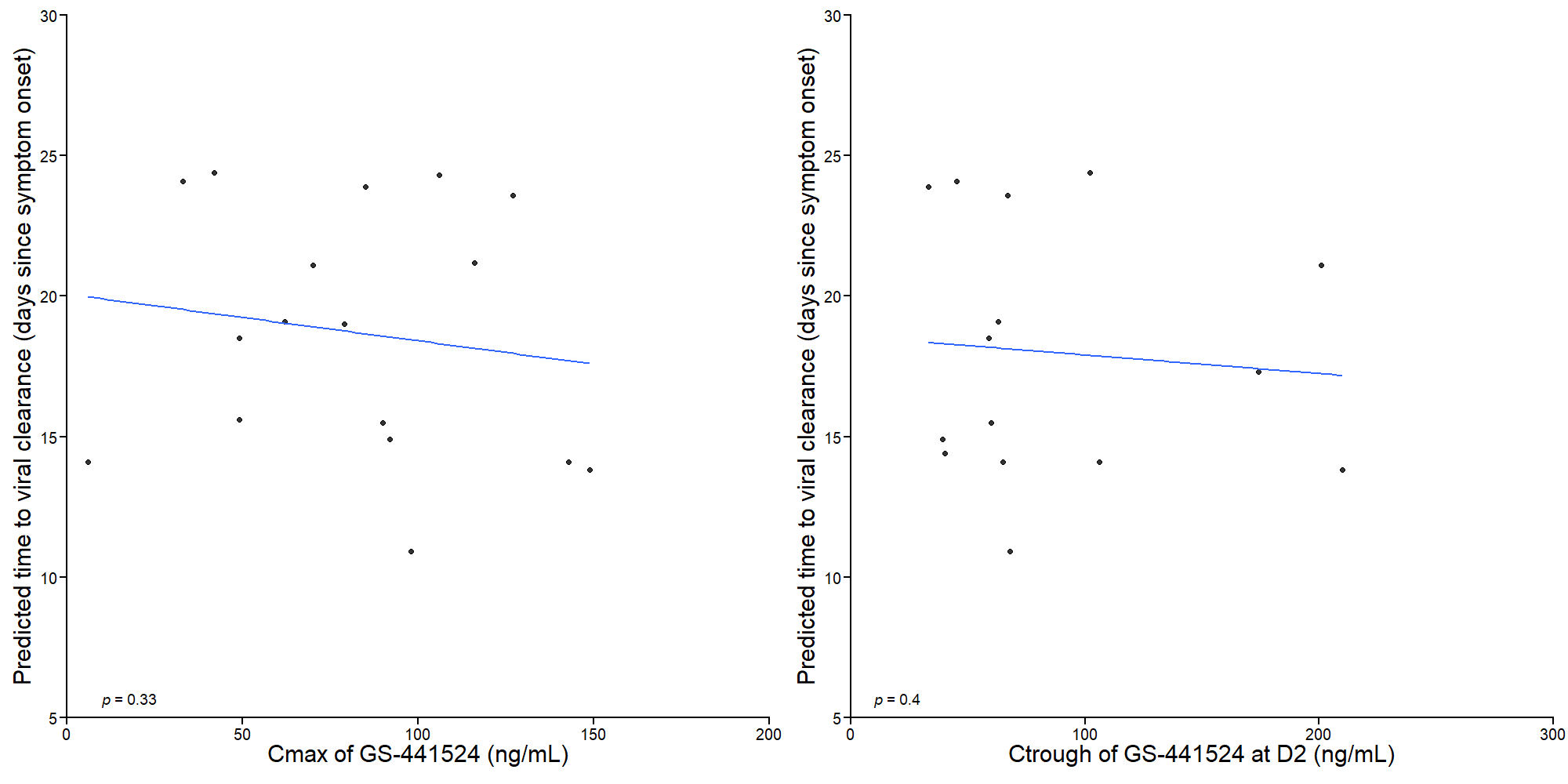

b)

a)

### Predicted individual time to viral clearance versus C_max_ at D1 (N=17, a) and C_trough_ at D2 (N=18, b) of GS-441524 in patients with viral load at baseline > 3.5 log_10_ copies/10^4^ cells

### Table S1. Population parameters of models including remdesivir efficacy starting between 0 and 5 days after randomization

| T_lag_ | 0 | | 1 | | 2 | | 3 | | 4 | | 5 | |
| --- | --- | --- | --- | --- | --- | --- | --- | --- | --- | --- | --- | --- |
|  | Fixed effect (RSE) | Random effect SD | Fixed effect (RSE) | Random effect SD | Fixed effect (RSE) | Random effect SD | Fixed effect (RSE) | Random effect SD | Fixed effect (RSE) | Random effect SD | Fixed effect (RSE) | Random effect SD |
| R_0_ | 10.2 (11.7) | $0.50$ | 10.0 (9.2) | 0.50 | 10.6 (10) | 0.50 | 10.7 (10.2) | 0.50 | 11.1 (10.4) | 0.50 | 10.8 (10.8) | 0.50 |
| $\boldsymbol{\delta}_{\mathbf{age}}\mathbf{(<}\mathbf{65}\mathbf{)}$ (d^-1^)  $\boldsymbol{\delta}_{\mathbf{age}}\boldsymbol{(\geq}\mathbf{65}\mathbf{)}$ (d^-1^) | 0.89 (4.5)  0.74 (29.2) | $0.45$  $-$ | 0.90 (4.2)  0.75 (32.0) | 0.45  $-$ | 0.88 (4.7)  0.73 (25) | 0.46  $-$ | 0.88 (4.5)  0.73 (30.1) | 0.46  $-$ | 0.88 (4.4)  0.73 (31.2) | 0.46  $-$ | 0.88 (4.4)  0.73 (31.0) | 0.45  $-$ |
| $\mathbf{p}$ (10^6^ virus. cells^-1^.d^-1^) | 1.30 (16.9) | $0.32$ | 1.4 (13.5) | 0.31 | 1.20 (25.0) | 0.40 | 1.20 (21.0) | 0.40 | 1.20 (19.3) | 0.34 | 1.20 (19.7) | 0.35 |
| $\boldsymbol{\varepsilon}$ (%) | **49 (27.0)**  **p-value=0.020** | $0.87$ | **50 (17.9) p-value=0.033** | 0.81 | **53 (13.3) p-value=0.0026** | 0.74 | **52 (16.1) p-value=0.0024** | 0.74 | **43 (35.2) p-value=0.19** | 1.36 | **37 (27.9) p-value=0.42** | 0.86 |
| $\boldsymbol{\sigma}$ (virus.mL^-1^) | 1.14 (2.27) | $-$ | 1.14 (2.18) | $-$ | 1.14 (2.18) | $-$ | 1.14 (2.28) | $-$ | 1.14 (2.19) | $-$ | 1.14 (2.30) | $-$ |

T_lag_: pharmacological delay between the administration and the beginning of antiviral efficacy; $R_{0}$: basic reproductive number; $\delta:$loss rate of infected cells; $p:$ rate of viral production; ɛ: remdesivir efficacy; σ: residual variability; RSE= relative standard error.

**Table S2a. Population parameters of models including remdesivir efficacy starting between 0 and 5 days after randomization in patients randomized ≤7 days since symptom onset**

| T_lag_ | 0 | | 1 | | 2 | | 3 | | 4 | | 5 | |
| --- | --- | --- | --- | --- | --- | --- | --- | --- | --- | --- | --- | --- |
|  | Fixed effect (RSE) | Random effect SD | Fixed effect (RSE) | Random effect SD | Fixed effect (RSE) | Random effect SD | Fixed effect (RSE) | Random effect SD | Fixed effect (RSE) | Random effect SD | Fixed effect (RSE) | Random effect SD |
| R_0_ | 17.3 (19.1) | $0.5$0 | 17.9 (20.6) | 0.50 | 18.4 (30.9) | 0.50 | 18.5 (39.6) | 0.50 | 17.6 (22.3) | 0.50 | 17.4 (20.6) | 0.50 |
| $\boldsymbol{\delta}_{\mathbf{age}}\mathbf{(<}\mathbf{65}\mathbf{)}$ (d^-1^)  $\boldsymbol{\delta}_{\mathbf{age}}\boldsymbol{(\geq}\mathbf{65}\mathbf{)}$ (d^-1^) | 0.92 (7.5)  0.84 (91.5) | 0.51  $-$ | 0.92 (7.5)  0.84 (91.5) | 0.53  $-$ | 0.90 (9.4)  0.81 (89) | 0.53  $-$ | 0.89 (11.4)  0.80 (90) | 0.53  $-$ | 0.91 (7.9)  0.82 (89.2) | 0.51  $-$ | 0.93 (7.4)  0.85 (90.5) | 0.51  $-$ |
| $\mathbf{p}$(10^6^ virus. cells^-1^.d^-1^) | 2.36 (25.5) | 0.24 | 2.43 (30.5) | 0.25 | 2.36 (20.9) | 0.25 | 2.16 (26.8) | 0.29 | 2.18 (33.8) | 0.24 | 2.48 (31.5) | 0.20 |
| $\boldsymbol{\varepsilon(\%)}$ | **12 (80.5) p-value=0.13** | 1.48 | **6 (97) p-value=0.16** | 1.21 | **43 (31.3) p-value=0.058** | 1.21 | **0.36 (49.7)**  **p-value=0.045** | 1.70 | **18 (57.8)**  **p-value=0.53** | 1.30 | **14 (51.8)**  **p-value=0.51** | 1.08 |
| $\boldsymbol{\sigma}$ (virus.mL^-1^) | 1.11 (3.4) | $-$ | 1.11 (3.4) | $-$ | 1.11 (3.4) | $-$ | 1.11 (3.2) | $-$ | 1.11 (3.5) | $-$ | 1.11 (3.4) | $-$ |

T_lag_: pharmacological delay between the administration and the beginning of antiviral efficacy; $R_{0}$: basic reproductive number; $\delta:$loss rate of infected cells; $p:$ rate of viral production; ɛ: remdesivir efficacy; σ: residual variability; RSE= relative standard error.

**Table S2b. Parameters distribution using model averaging (Median, 95% CI) in patients randomized ≤7 days since symptom onset**

|  | Parameter estimates |  |
| --- | --- | --- |
| **Parameter** | **Fixed effects (Median. 95% CI)** | **Random effect SD (Median. 95% CI)** |
| **R_0_** | 18.09 (6.29-30.26) | 0.5 |
| $\boldsymbol{\delta}_{\mathbf{age}}\mathbf{(<}\mathbf{65}\mathbf{)}$ **(d^-1^)**  $\boldsymbol{\delta}_{\mathbf{age}}\boldsymbol{(\geq}\mathbf{65}\mathbf{)}$ **(d^-1^)** | 0.90 (0.72-1.07)  0.82 (0.66-0.98) | 0.52 (0.44-0.60) |
| $\mathbf{p}$ **(10^6^ virus.cell^-1^.d^-1^)** | 2.20 (1.10-3.40) | 1.43 (0.24-3.07) |
| $\boldsymbol{\varepsilon}$**(%)** | 30 (-0.2-68) | 0.55 (-0.61-1.85) |

$R_{0}$: basic reproductive number; $\delta:$loss rate of infected cells; $p:$ rate of viral production; ɛ: remdesivir efficacy

**Table S3a. Population parameters of models including remdesivir efficacy starting between 0 and 5 days after randomization, in patients randomized** >**7 days since symptom onset**

| T_lag_ | 0 | | 1 | | 2 | | 3 | | 4 | | 5 | |
| --- | --- | --- | --- | --- | --- | --- | --- | --- | --- | --- | --- | --- |
|  | Fixed effect (RSE) | Random effect SD | Fixed effect (RSE) | Random effect SD | Fixed effect (RSE) | Random effect SD | Fixed effect (RSE) | Random effect SD | Fixed effect (RSE) | Random effect SD | Fixed effect (RSE) | Random effect SD |
| R_0_ | 6.74 (18.3) | 0.50 | 6.76 (14.7) | 0.50 | 6.92 (9.8) | 0.50 | 6.98 (9.8) | 0.50 | 7.77 (9.51) | 0.50 | 7.50 (9.34) | 0.50 |
| $\boldsymbol{\delta}_{\mathbf{age}}\mathbf{(<}\mathbf{65}\mathbf{)}$ (d^-1^)  $\boldsymbol{\delta}_{\mathbf{age}}\boldsymbol{(\geq}\mathbf{65}\mathbf{)}$ (d^-1^) | $0.94 (5.6)$  $0.77 (84.5)$ | $0.35$  $-$ | 0.94 (5.5)  0.77 (25.8) | 0.35  $-$ | 0.94 (5.3)  0.77 (25.4) | 0.34  $-$ | 0.94 (5.2)  0.76 (24.9) | 0.34  $-$ | 0.92 (4.9)  0.75 (25.4) | 0.36  $-$ | 0.93 (5.0)  0.76 (24.5) | 0.35  $-$ |
| $\mathbf{p}$ (10^5^ virus. cells^-1^. d^-1^) | $6.66 (84.5)$ | $0.63$ | 7.50 (60) | 0.60 | 6.80 (30.1) | 0.62 | 7.00 (31.3) | 0.60 | 8.70 (20.8) | 0.48 | 8.82 (25.1) | 0.47 |
| $\boldsymbol{\varepsilon}$ (%) | **46 (59.6) p-value=0.9** | $0.85$ | **52 (27.9) p-value=0.9** | 0.73 | **49 (20.7) p-value=0.9** | 0.63 | **51 (20) p-value=0.7** | 0.61 | **0.41 (47.1) p-value=0.8** | 0.88 | **41 (24.5) p-value=0.7** | 0.61 |
| $\boldsymbol{\sigma}$ (virus.mL^-1^) | $1.14 (2.8)$ | $-$ | $1.14 (2.8)$ | $-$ | $1.14 (2.8)$ | $-$ | $1.14 (2.8)$ | $-$ | $1.14 (2.8)$ | $-$ | $1.14 (2.8)$ | $-$ |

T_lag_: pharmacological delay between the administration and the beginning of antiviral efficacy; $R_{0}$: basic reproductive number; $\delta:$loss rate of infected cells; $p:$ rate of viral production; ɛ: remdesivir efficacy; σ: residual variability; RSE= relative standard error.

**Table S3b. Parameters distribution using model averaging (Median, 95% CI) in patients randomized >7 days since symptom onset**

|  | Parameter estimates |  |
| --- | --- | --- |
| **Parameter** | **Fixed effects (Median. 95% CI)** | **Random effect SD (Median. 95% CI)** |
| **R_0_** | 6.91 (4.97-8.65) | 0.5 |
| $\boldsymbol{\delta}_{\mathbf{age}}\mathbf{(<}\mathbf{65}\mathbf{)}$ **(d^-1^)**  $\boldsymbol{\delta}_{\mathbf{age}}\boldsymbol{(\geq}\mathbf{65}\mathbf{)}$ **(d^-1^)** | 0.94 (0.84-1.04)  0.77 (0.68-0.87) | 0.34 (0.28-0.41) |
| $\mathbf{p}$ **(10^5^ virus.cell^-1^.d^-1^)** | 7.1 (-0.7-15) | 0.61 (0.03-1.19) |
| $\boldsymbol{\varepsilon}$**(%)** | 50 (16-82) | 0.69 (0.05-1.41) |

$R_{0}$: basic reproductive number; $\delta:$loss rate of infected cells; $p:$ rate of viral production; ɛ: remdesivir efficacy

### Table S4a. Parameters distribution using model averaging (Median, 95% CI) in patients with viral load at admission$\boldsymbol{\geq}$3.5 log_10_ copies/10^4^ cells

|  | Parameter estimates |  |
| --- | --- | --- |
| **Parameter** | **Fixed effects (Median. 95% CI)** | **Random effect SD (Median. 95% CI)** |
| **R_0_** | 8.68 (6.59-10.75) | 0.5 |
| $\boldsymbol{\delta}_{\mathbf{age}}\mathbf{(<}\mathbf{65}\mathbf{)}$ **(d^-1^)**  $\boldsymbol{\delta}_{\mathbf{age}}\boldsymbol{(\geq}\mathbf{65}\mathbf{)}$ **(d^-1^)** | 0.70 (0.63-0.77)  0.65 (0.55-0.75) | 0.21 (0.15-0.27) |
| $\mathbf{p}$ **(10^6^ virus.cell^-1^.d^-1^)** | 3.51 (2.80-4.20) | 0.11 (0.02-0.21) |
| $\boldsymbol{\varepsilon}$**(%)** | 80 (64-96) | 2.17 (0.67-3.73) |

$R_{0}$: basic reproductive number; $\delta:$loss rate of infected cells; $p:$ rate of viral production; ɛ: remdesivir efficacy

**Table S4b. Population parameters of models including remdesivir efficacy starting between 0 and 5 days after randomization in patients with viral load at admission** $\boldsymbol{\geq}$**3.5 log_10_ copies/10^4^ cells**

| T_lag_ | 0 | | 1 | | 2 | | 3 | | 4 | | 5 | |
| --- | --- | --- | --- | --- | --- | --- | --- | --- | --- | --- | --- | --- |
|  | Fixed effect (RSE) | Random effect SD | Fixed effect (RSE) | Random effect SD | Fixed effect (RSE) | Random effect SD | Fixed effect (RSE) | Random effect SD | Fixed effect (RSE) | Random effect SD | Fixed effect (RSE) | Random effect SD |
| R_0_ | 8.66 (11.6) | 0.50 | 8.68 (13) | 0.50 | 9.63 (12.8) | 0.50 | 9.94 (11.5) | 0.50 | 9.46 (11.7) | 0.50 | 9.51 (11.6) | 0.50 |
| $\boldsymbol{\delta}_{\mathbf{age}}\mathbf{(<}\mathbf{65}\mathbf{)}$ (d^-1^)  $\boldsymbol{\delta}_{\mathbf{age}}\boldsymbol{(\geq}\mathbf{65}\mathbf{)}$ (d^-1^) | 0.70 (5.2)  0.65 (63.6) | 0.21  $-$ | 0.70 (5.3)  0.65 (60.5) | 0.21  $-$ | 0.71 (6.6)  0.65 (54.3) | 0.22  $-$ | 0.71 (5.0)  0.65 (55) | 0.23  $-$ | 0.74 (5.0)  0.67 (58.6) | 0.23  $-$ | 0.74 (5.36)  0.68 (61.9) | 0.23  $-$ |
| $\mathbf{p}$ (10^6^ virus. cells^-1^.d^-1^) | 3.56 (7.7) | 0.11 | 3.49 (12.1) | 0.12 | 3.36 (20.6) | 0.13 | 3.50 (8.9) | 0.10 | 3.72 (10) | 0.08 | 3.53 (8.5) | 0.09 |
| $\boldsymbol{\varepsilon(\%)}$ | **80 (10.4) p-value<10^-5^** | 2.16 | **82 (9.8)**  **p-value<10^-5^** | 2.22 | **50 (62.1)**  **p-value<10^-4^** | 3.33 | **55 (43)**  **p-value=10^-4^** | 3.09 | **18 (78)**  **p-value=0.59** | 2.47 | **10 (128)**  **p-value=0.81** | 2.64 |
| $\boldsymbol{\sigma}$ (virus.mL^-1^) | 1.09 (3.3) | $-$ | 1.08 (3.4) | $-$ | 1.10 (3.4) | $-$ | 1.10 (3.3) | $-$ | 1.12 (3.3) | $-$ | 1.12 (3.3) | $-$ |

T_lag_: pharmacological delay between the administration and the beginning of antiviral efficacy; $R_{0}$: basic reproductive number; $\delta:$loss rate of infected cells; $p:$ rate of viral production; ɛ: remdesivir efficacy; σ: residual variability; RSE= relative standard error.

### Table S5a. Parameters distribution using model averaging (Median, 95% CI) in patients with viral load at admission < 3.5 log_10_ copies/10^4^ cells

|  | Parameter estimates |  |
| --- | --- | --- |
| **Parameter** | **Fixed effects (Median. 95% CI)** | **Random effect SD (Median. 95% CI)** |
| **R_0_** | 55.48 (11.31-110.70) | 0.5 |
| $\boldsymbol{\delta}_{\mathbf{age}}\mathbf{(<}\mathbf{65}\mathbf{)}$ **(d^-1^)**  $\boldsymbol{\delta}_{\mathbf{age}}\boldsymbol{(\geq}\mathbf{65}\mathbf{)}$ **(d^-1^)** | 0.49 (0.40-0.58)  0.48 (0.33-0.64) | 0.47 (0.38-0.55) |
| $\mathbf{p}$ **(10^4^ virus.cell^-1^.d^-1^)** | 1.26 (0.025-2.61) | 0.56 (-0.013-1.37) |
| $\boldsymbol{\varepsilon}$**(%)** | 64 (0-1) | 2.35 (-0.42-7.47) |

$R_{0}$: basic reproductive number; $\delta:$loss rate of infected cells; $p:$ rate of viral production; ɛ: remdesivir efficacy

**Table S5b. Population parameters of models including remdesivir efficacy starting between 0 and 5 days after randomization, in patients with viral load at admission < 3.5 log_10_ copies/10^4^ cells**

| T_lag_ | 0 | | 1 | | 2 | | 3 | | 4 | | 5 | |
| --- | --- | --- | --- | --- | --- | --- | --- | --- | --- | --- | --- | --- |
|  | Fixed effect (RSE) | Random effect SD | Fixed effect (RSE) | Random effect SD | Fixed effect (RSE) | Random effect SD | Fixed effect (RSE) | Random effect SD | Fixed effect (RSE) | Random effect SD | Fixed effect (RSE) | Random effect SD |
| R_0_ | $55.3 (40.1)$ | $0.50$ | 60.7 (60.1) | 0.50 | 51.2 (43.1) | 0.50 | 65.6 (92.1) | 0.50 | 48.7 (36.9) | 0.50 | 64.2 (35.3) | 0.50 |
| $\boldsymbol{\delta}_{\mathbf{age}}\mathbf{(<}\mathbf{65}\mathbf{)}$ (d^-1^)  $\boldsymbol{\delta}_{\mathbf{age}}\boldsymbol{(\geq}\mathbf{65}\mathbf{)}$ (d^-1^) | $0.53 (8.40)$  $0.52 (640)$ | $0.43$  $-$ | 0.52 (9.6)  0.51 (672) | 0.43  $-$ | 0.51 (8.73)  0.50 (823) | 0.44  $-$ | 0.49 (9.36)  0.48 (851) | 0.45  $-$ | 0.49 (7.98)  0.48 (318) | 0.46  $-$ | 0.48 (7.36)  0.47 (772) | 0.46  $-$ |
| p (10^4^ virus. cells^-1^.d^-1^) | $1.93 (40.0)$ | $0.58$ | 2.1 (45.4) | 0.74 | 1.53 (43.4) | 0.79 | 1.48 (49.5) | 0.57 | 1.17 (42.5) | 0.44 | 1.34 (58.5) | 0.44 |
| $\boldsymbol{\varepsilon}$ (%) | **10 (77.8) p-value=1** | $0.60$ | **11 (67.4) p-value=1** | 0.34 | **30 (74.8) p-value=0.25** | 0.60 | **49 (88.2) p-value=0.08** | 1.09 | **67 (31.7) p-value=0.013** | 3.92 | **62 (61.4) p-value=0.014** | 6.22 |
| $\boldsymbol{\sigma}$ (virus.mL^-1^) | $0.97 (3.6)$ | $-$ | $0.97 (3.6)$ | $-$ | $0.97 (3.6)$ | $-$ | $0.97 (3.6)$ | $-$ | $0.97 (3.6)$ | $-$ | $0.97 (3.6)$ | $-$ |

T_lag_: pharmacological delay between the administration and the beginning of antiviral efficacy; $R_{0}$: basic reproductive number; $\delta:$loss rate of infected cells; $p:$ rate of viral production; ɛ: remdesivir efficacy; σ: residual variability; RSE= relative standard error.

| Characteristics | Patients with LRT samples (N=120) |
| --- | --- |
| Male gender* | 88 (72.7%) |
| Age*  Age <65  Age ≥65 | 67.5 (59-73)  46 (38.3%)  74 (61.7%) |
| Admitted in Intensive Care Units | 105 (87.5%) |

**Table S6. Characteristics of the population with in lower respiratory tract samples**
